# Study of *PLEKHA1* gene polymorphism and their association with age related macular degeneration in Indian patients

**DOI:** 10.1101/2021.05.05.21255960

**Authors:** Divya Gupta, Shobhit Chawla, Shubha R. Phadke

## Abstract

**Background:** Age-related macular degeneration (AMD) is an important cause of visual impairment in elderly people. AMD is a multifactorial disease in which both environmental and genetic factors have been implicated. Various single nucleotide polymorphisms (SNPs) have been found to be associated with AMD.

**Aim:** This study was aimed to investigate the association of polymorphisms in *PLEKHA1* rs4146894 (G>A) gene with age related macular degeneration (AMD) in Indian patients.

**Method:** Genotyping for the *PLEKHA1* rs4146894 (G>A) polymorphism performed in 121 AMD patients and 100 controls by polymerase chain reaction (PCR) and sequencing method.

**Results:** This is the first study from India on *PLEKHA1* rs4146894 (G>A) polymorphism. In this study, we did not get any change in DNA sequence by sequencing method. So, we can say that the risk allele of *PLEKHA1* rs4146894 (G>A) polymorphism is not find in Indian population. On the other hand in Indian population the risk of A allele is not present, and this gene is not playing any significant contribution with AMD in Indian population.

**Conclusion:** Very limited literatures are available for *PLEKHA1* polymorphism in AMD patients, though, it is quite possible that the variation in the data shows population based differences. So, the present study raises the possibility that *PLEKHA1* rs4146894 (G>A) polymorphism may not have a significant association with AMD in Indian patients.

## INTRODUCTION

Age-related macular degeneration (ARMD or AMD; OMIM 603075) is the degenerative disease of retina that causes progressive impairment of central vision leading to irreversible vision loss in elderly people. Age-related macular degeneration (AMD) affects the macular region of the retina. The macula has a high density of photoreceptors which provides detailed central vision. In early stage it is referred to as age-related maculopathy (ARM). The prevalence of the disease increases with age, affecting 9% of the population over age 65 years and 30% over age 70 years [1]. Fewer than 1% of patients are less than 65 years of age. In India, the prevalence of AMD ranges 1.84%–2.7% in different epidemiological studies [2]. In the United States alone, more than 8 million have intermediate AMD and nearly 2 million have advanced AMD. It is estimated that approximately 80 milion people will be affected due to retinal complications including AMD by the year 2020 [3]. The etiology of AMD is largely unknown and is generally considered to be a complex multifactorial disorder with significant contribution of genetics factors. Though both environment and genetic factors are implicated; its actual etiology is largely unknown. Polymorphisms in *CFH, ARMS2* and *HTRA1* have been consistently shown to be associated with AMD across different populations. In addition, other genes like *ApoE, CX3CR1, PLEKHA1* and *VEGF* were studied for their association with AMD. Neovascularization is an important component of pathology of AMD.

Whole genome association study in familial cases with AMD identified a linkage peak for AMD at 10q26, most highly significant between a *PLEKHA1* involved in phospho-inositide metabolism and *ARMS2* [4,5]. *PLEKHA1* rs4146894 G>A may play independent and possibly synergistic role for development of AMD. Due to limited information about many genes and especially from Indian populations, the present study was planned.

## MATERIALS AND METHODS

### Patients and Clinical Examination

The study protocol was approved by the Ethics Committee of Sanjay Gandhi Post Graduate Institute of Medical Sciences Raibareli Road Lucknow, Uttar Pradesh. All participants received a standard ophthalmological examination, including visual acuity measurement, slit-lamp biomicroscopy and dilated fundus examination performed by a retinal specialist. The diagnosis was confirmed in fluorescein angiography and cases with both; dry and wet types of AMD were included. Controls were more than 70 years of age and were confirmed not to have clinical evidence of AMD in any of the eyes by opthalmological examination. Sample size was calculated before the study with the use of Quanto software version 1.2 by institutional statistician. The variables set were as follows-minimum allele frequency of gene 0.30, relative risk 2, prevalence 0.1% and type one error (alpha) as 0.05. For the 80% power of the study, the sample size was calculated to be 92. Samples were collected after obtaining informed consent from 121 clinically diagnosed AMD cases and 100 controls. There remained no conflict of interest pertaining to this study.

### DNA preparation and genotyping

Five milliliter of peripheral venous blood was collected in EDTA vacationer from all participants and stored at -80ºC. Genomic DNA was extracted by using commercially available genomic DNA extraction kit QIAamp DNA extraction Kit (Qiagen Blood DNA Mini Kit, Hilden, Germany), according to the manufacturer’s instructions. Genotyping was performed by polymerase chain reaction and Sequencing method. For *PLEKHA1* a 393bp fragment was amplified from the genomic DNA using primers Forward 5’-GGATCACTTTGGGGCTTCC -3’ Reverse 5’-GCCTTTGTGGAAGGAAGACA -3’ which correspond to the intronic region. The reaction mixture of *PLEKHA1* polymorphism consisted of: 10X buffer with MgCl_2_ 2.5 μl, Taq 3 U/ μl-0.5 μl, dNTP 10 milimoles - 1 μl, forward primer 10 picomoles - 1μl, Reverse primer 10 picomoles - 1 μl, DNA – 1.5 μl (50 ng/μl) and HPLC water 17.5 μl per 25μl. PCR conditions were as follows: hot lid 105ºC for 5 min, denaturation 95ºC for 5 min, 38 cycles consisting of denaturation at 94 ºC for 45 seconds, 68.0 ºC annealing for 45 seconds and extension at 72 ºC for 30 seconds, followed by final extension at 72 ºC for 5 minutes and hold at 15 ºC forever. The amplified products were tested on 2% agarose gels pre-stained with ethidium bromide with a migrating distance of approximately 3 cm, and the product bands were visualized under ultraviolet lights, samples were then used for SBT using ABI 310 sequence analyzer (Applied Biosystems Corporation, CA, USA) showed in supplementary figure 1.

### Statistical Analysis

In this study, we find only homozygous wild genotype (GG). So, we did not apply any statistical analysis for the *PLEKHA1* rs4146894 (G>A) polymorphism.

## RESULTS

A total of 121 patients with AMD (males-74 & females-47) and 100 controls (males-71 & females-29) without AMD were recruited after detailed ophthalmological evaluation. The mean age was 70.40 (60 – 96) years for AMD patients and 74.42 (70-95) years for controls. Genotypes were determined successfully by PCR, and sequencing methods. This is the first study from India on Indian AMD samples to analysis the *PLEKHA1* rs4146894 (G>A) polymorphism. After completing of genotyping analysis of AMD patients and control samples, we did not get any genotypic changes in the *PLEKHA1* rs4146894 G>A. In all samples of AMD patients and as well as in controls, sequences of *PLEKHA1* rs4146894 gene were GG genotype showed in Supplementary Figure 1.

## DISCUSSION

Understanding about etiology of AMD is yet limited. Whole genome wide association showed linkage peak for AMD at 10q26 where *PLEKHA1* and LOC387715 (ARMS2) genes are located and both have shown independent association with AMD [6,7]. A study from French population by Nicolas et al., showed the polymorphism rs4146894 G>A was associated with AMD their odds ratio for the mutant homozygous genotype (AA) was 9.1; p-value less than or equal to 0.0001, and for the heterozygous genotype (GA) was 2.6; p-value less than or equal to 0.04. The odds ratio for mutant allele (A) frequency was 0.67 in exudative AMD patients and 0.41 in controls with the p-value less than or equal to 0.0001 [8]. However in the present study did not find association with *PLEKHA1* rs4146894 G>A on an Indian AMD patients.

Available limited data about *PLEKHA1* shows an association with the AMD; whereas the present study did not show any association of with *PLEKHA1* rs4146894 G>A with AMD in an Indian population. Further studies in Indian populations are needed to confirm these findings in larger groups of patients.

## CONCLUSION

Very limited literatures are available for *PLEKHA1* polymorphism in AMD patients, though, it is quite possible that the variation in the data shows population based differences. So, the present study raises the possibility that *PLEKHA1* rs4146894 (G>A) polymorphism may not have a significant association with AMD in Indian patients.

## Data Availability

All data presented in this manuscript is authentic.

## DECLARATION

This work has not been published anywhere previously and that it is not simultaneously being considered for any other publication and I am responsible for the integrity and accuracy of the data analysis.

### Ethical Approval and Consent to Participate

The study protocol was approved by the Ethics Committee of Sanjay Gandhi Post Graduate Institute of Medical Sciences, Lucknow, Uttar Pradesh (IEC Code No: A-03:PGI/SRF/IEC/54/29.4.2011)

### Consent for Publication

All authors approve the manuscript for publication.

### Availability of Data and Materials

All authors approve the manuscript for publication.

### Author’s Contribution

Divya Gupta made effort on study design, collection of samples, complete the bench work, literature search, manuscript writing, data analysis. Shobhit Chawla provided the sample for the study and made the diagnosis confirmed. Shubha R. Phadke made effort on study design, provided laboratory support and correction in manuscript.

## Acknowledgement

I would like to express my greatest appreciation to all the participants in the study. This work was supported by the Indian Council of Medical Research (Grant No. 45/20/2010/CMB/BMS) New Delhi to Divya Gupta, award of ICMR-SRF (Indian Council of Medical Research-Senior Research Fellow).

---

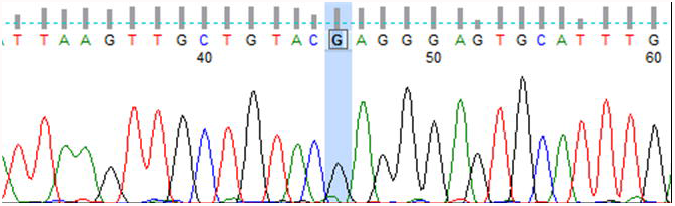

